# Movement-responsive deep brain stimulation for Parkinson’s Disease using a remotely optimized neural decoder

**DOI:** 10.1101/2024.08.14.24312002

**Authors:** Tanner C. Dixon, Gabrielle Strandquist, Alicia Zeng, Tomek Fraczek, Raphael Bechtold, Daryl Lawrence, Shravanan Ravi, Philip A. Starr, Jack L. Gallant, Jeffrey A. Herron, Simon J. Little

## Abstract

Deep brain stimulation (DBS) has garnered widespread use as an effective treatment for advanced Parkinson’s Disease (PD). Conventional DBS (cDBS) provides electrical stimulation to the basal ganglia at fixed amplitude and frequency, yet patients’ therapeutic needs are often dynamic with residual symptom fluctuations or side-effects. Adaptive DBS (aDBS) is an emerging technology that modulates stimulation with respect to real-time clinical, physiological, or behavioral states, enabling therapy to dynamically align with patient-specific symptoms. Here, we report a novel aDBS algorithm intended to mitigate movement slowness by delivering targeted stimulation increases during movement using decoded motor signals from the brain. Compared to cDBS and a control algorithm that decreases stimulation during movement, our approach demonstrates enhanced clinical efficacy, with improvements in movement speed, reduced dyskinesia, and better patient-reported therapeutic quality. Furthermore, we demonstrate proof-of-principle of a machine learning pipeline capable of remotely optimizing aDBS parameters in a home setting. This work illustrates the potential of movement-responsive aDBS as a new therapeutic approach and highlights how machine learning assisted programming can simplify optimization to facilitate translational scalability.

## INTRODUCTION

Deep brain stimulation (DBS) has become a widely adopted treatment for patients with advanced Parkinson’s Disease (PD) and medication refractory symptoms. Conventional DBS (cDBS) delivers constant electrical pulses to the basal ganglia at fixed amplitudes and frequencies. While the exact mechanisms are still being elucidated (Ashkan et al., 2017; McGregor & Nelson, 2019), cDBS has been shown to alleviate movement slowness (bradykinesia, the cardinal symptom of PD) and tremor, while reducing involuntary movements (dyskinesia) that arise as a side-effect of dopaminergic medication (Deuschl et al., 2006; Follett et al., 2010; Odekerken et al., 2013). However, residual fluctuations due to medication and activity/arousal levels often persist (Pozzi & Isaias, 2022), as the static nature of cDBS fails to address these dynamic clinical needs. This can create challenging trade-offs between maximizing clinical efficacy and minimizing side-effects when programming cDBS therapy.

Adaptive DBS (aDBS) seeks to overcome the limitations of cDBS by modulating stimulation in accordance with real-time clinical, physiological, or behavioral states. These states can be tracked through local field potential (LFP) biomarkers recorded from the brain. Beta-band (13-30Hz) LFP activity in the basal ganglia, which covaries with PD symptom severity and medication state (Kühn et al., 2006; Lofredi et al., 2023; Neumann et al., 2016), has been used as the control signal in the majority of aDBS algorithms previously tested (Little & Brown, 2020). The earliest approaches to be developed treated beta as a trigger for on-off stimulation control, targeting sub-second bursts of activity with putative roles in pathological brain circuit dynamics (He et al., 2023; Little et al., 2013, 2016; Tinkhauser et al., 2017). These studies using “fast” algorithms have provided collective evidence that aDBS may increase efficacy and reduce side-effects through more selective stimulation. Later approaches have begun considering longer timescales and proportional control algorithms more suitable for counteracting the gradual fluctuations of concurrent medication cycles (Arlotti et al., 2018; Gilron, Little, Perrone, et al., 2021; Rosa et al., 2017; Velisar et al., 2019). Both fast and slow beta algorithms are currently in multi-centre clinical trials [ADAPT-PD: NCT04547712]. Notably, however, this same beta signal is reduced during movement (Eisinger et al., 2020; Kühn et al., 2004). This raises concern that beta-driven aDBS will have the unintended consequence of reducing stimulation during movement (He et al., 2023; Iturrate et al., 2019; Johnson et al., 2016; Little & Brown, 2020), which is precisely when the prokinetic effects of DBS are theoretically most necessary. Broader exploration of biomarkers and novel algorithm design will be required to navigate these problematic interactions between symptoms, biomarkers, and movement.

While most aDBS algorithms attempt to identify latent clinical states that reflect probability of symptoms, an alternative approach would be to target movement itself. Bradykinesia in PD characteristically involves difficulty initiating voluntary movement with progressive reduction in vigor during repetitive action (Bologna et al., 2020). We therefore sought to develop a movement-responsive aDBS algorithm that selectively increases stimulation during movement for treating bradykinesia, while reducing stimulation during rest to mitigate dyskinesia (Figure 1A-B). The gating theory of basal ganglia function in PD pathophysiology (Chevalier & Deniau, 1990; Ivry & Spencer, 2004) provides a theoretical mechanism of action supporting this approach. Under healthy circumstances, the basal ganglia inhibit movement until an action has been selected and released (Klaus et al., 2019). In PD, excessive inhibition prevents release, leading to slow, difficult-to-initiate movement. By decoding motor intent to guide targeted stimulation, movement-responsive aDBS would selectively disinhibit movement when it is intended. This approach has been tested in a non-human primate model of PD using cortical gamma to indicate movement, showing therapeutic benefits comparable to conventional stimulation while reducing overall current delivery (Darbin et al., 2022). Movement- and postural-state dependent aDBS has also been explored for essential tremor with either cortical (Ferleger et al., 2020; Herron et al., 2017; Opri et al., 2020) or thalamic (He et al., 2021) algorithm inputs. Furthermore, selectively stimulating the indirect pathway during periods of increased movement has been shown to potentiate faster movements (Yttri & Dudman, 2016). This suggests that movement-responsive DBS could also facilitate beneficial motor network learning and plasticity that facilitates improved movements over time.

**Figure 1.**
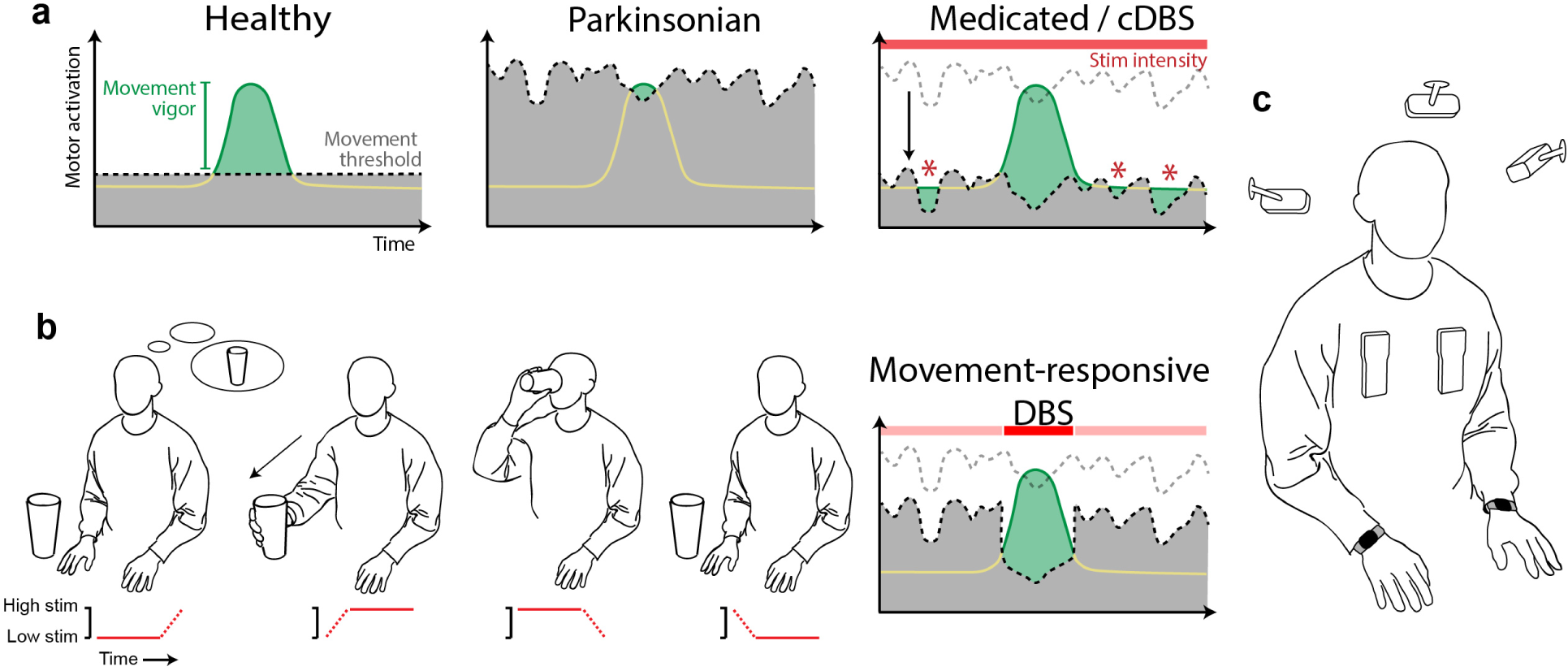
Movement-responsive DBS theoretical framework. (a) In the neurologically healthy state (left) the basal ganglia inhibit actions that have not yet been selected through activation beyond a gating threshold (black dashed line). When activation associated with a particular action reaches this threshold, movement is released with the appropriate timing and vigor (subthreshold activation is represented as a yellow line, which becomes green to indicate suprathreshold activation/selection and downstream movement). In the Parkinsonian state (middle), excessive and fluctuating inhibition in the basal ganglia raises the movement threshold. This results in difficulty initiating vigorous movements. In the treated Parkinsonian state (Medicated/cDBS, right), the movement threshold is lowered through tonic disinhibition without regard for motor intent. This can result in unintended movements (red asterisks) when activation exceeds the reduced, fluctuating threshold. (b) A “Movement-responsive” framework is proposed where stimulation is tied to motor intent, reaching higher levels during intended movement and reducing to lower levels at rest. The far right plot illustrates how this prevents dyskinesia when movement is not intended while providing targeted disinhibition for vigorous, volitional movement (compare to plots in panel (a)). (c) To enable movement-responsive experiments, neural data were streamed from bilateral STN and cortical electrodes in a participant implanted with the Medtronic Summit RC+S DBS system. External devices for streaming data from the implants were worn over the upper chest; however, adaptive stimulation was performed embedded on the device. Video from three cameras and wrist-worn accelerometry (Apple Watches) were simultaneously recorded and time-synced (note that the camera placement in the cartoon image is for illustration only and not indicative of the precise locations and orientations used in the study).

In the present study, we report the first fully embedded movement-responsive aDBS in a human subject with PD. We first optimized neural decoders using a custom machine learning pipeline seeking to predict movement from cortical and subthalamic LFP’s. Algorithm tuning was performed in a data-driven, automated fashion using data collected independently by the patient in their own home without any direct researcher involvement. We then assessed offline performance to evaluate the pipeline, in particular the benefit associated with including cortical signals and using a custom power band personalization method. Finally, we used the optimized algorithms to drive movement-responsive aDBS and compared it to conventional DBS and a control aDBS condition, demonstrating comparable or superior efficacy across all categories of motor evaluation, including movement speed, dyskinesia, and subjective patient reports of therapeutic quality.

## RESULTS

### Embedded aDBS functionality and algorithm tuning overview

One subject with PD who had previously received bilateral DBS implants with embedded adaptive capabilities (Medtronic RC+S, Figure 1C) was recruited for the study. The hardware in each hemisphere consisted of a depth lead for stimulating and sensing in the subthalamic nucleus (STN), a sensorimotor electrocorticography (ECoG) strip for sensing over the precentral (PreC, motor) and postcentral (PostC, somatosensory) cortices, and a neurostimulator for driving adaptive stimulation using LFP’s from the three sensed brain areas. The onboard algorithm guiding stimulation used linear discriminant analysis with additional layered operations for controlling the state transition dynamics (Sellers et al., 2021). LFP’s were first transformed into power bands with programmable frequency limits and smoothing properties, then up to four power bands were linearly combined and passed through a series of threshold-based logic operations to define the algorithm state. Our aim was to optimize the algorithm parameters in each stimulator for decoding movement of the contralateral arms and delivering targeted stimulation increases during their respective movements (Figure 1A-B).

A remote data collection platform (Strandquist et al., 2023) was deployed in the participant’s home office to collect model training data and evaluate algorithm performance. In addition to neural data, we recorded accelerometry from bilateral Apple Watches (thresholded to label as moving or stationary), multi-view video recordings, and keylogging from the participant’s home computer (Figure 1C). Five days of data were collected for model training and validation, while a sixth day was dedicated for model testing. Each recording day lasted approximately 25 minutes, during which the participant performed a self-guided series of clinically-standardized movement tasks using both hands (Unified Parkinson’s Disease Rating Scale Part III: rest tremor, finger tapping, hand movements - open-close, pronation-supination) as well as a typing task (Figure 2A,B).

**Figure 2.**
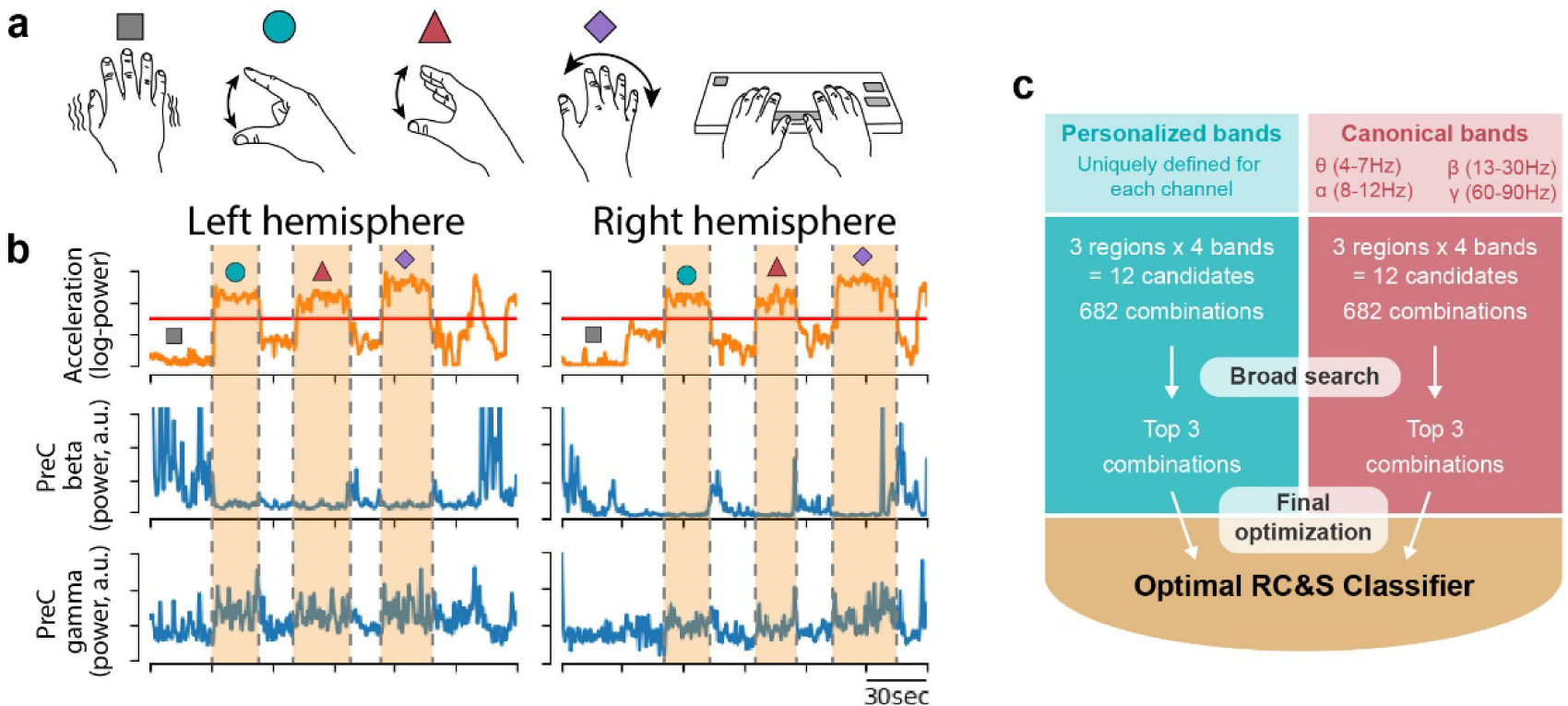
Optimizing the RC&S movement classifier. (a) Six days of data were collected for model training, testing, and validation. During each session the participant performed a series of self-guided clinical tasks: rest recording (assessing tremor/dyskinesia), rhythmic finger tapping, hand open-close, wrist pronation-supination, and typing (pictured in order from left to right). The participant alternated between hands such that one hand was always resting while the other performed the tasks, except for when typing bimanually. (b) Wrist-worn accelerometry was recorded during behavior, and LFP’s were simultaneously recorded from three brain regions: the STN and the Postcentral and Precentral gyri. A three-minute example window is displayed showing contralateral wrist acceleration alongside beta (13-30Hz) and gamma (60-90Hz) power bands recorded from the PreCentral gyrus (PreC). A threshold was applied to the wrist accelerometry (red line) to label binary moving and non-moving states. Movement periods are indicated by orange shading, with movement tasks indicated by symbols corresponding to the cartoons in (a). (c) An automated pipeline for feature selection and parameter optimization was designed for programming the RC+S to optimally classify movement state. Prior to parameter optimization, two sets of candidate power bands were identified that would later be used as stimulation control signals: “Personalized” bands, computed using a modified form of principal components decomposition, and “Canonical” bands, defined by traditional frequency ranges used in existing literature. A progressive narrowing of each candidate pool began with creating separate models for every possible combination of power bands and feeding each through a brief (20 iteration) Bayesian optimization routine (“Broad search”). The top three models underwent an additional 200 iterations in a “Final optimization” stage, after which the single top performer was selected for each hemisphere.

Given the unique architecture of the neurostimulators’ algorithms, a custom machine learning pipeline was developed for optimization (Figure 2C). In brief, we first fit coefficient weights for predicting continuous-valued watch accelerometry from neural inputs. Using Bayesian optimization, we then tuned the parameters that define the algorithm dynamics and translate the accelerometry predictions into binary movement state predictions. Finally, we applied a feature selection routine to search for the optimal set of neural inputs to the algorithm.

### Personalizing neural inputs improves algorithm performance

Since power band inputs may be defined over any frequency range, a method of constraining the search space was required. We used an approach that first identified candidate pools of potentially information-rich power bands, then progressively narrowed the combinatorial field to a single set of optimal inputs. We compared two methods of identifying candidates. The first method selected “canonical” power bands, representing commonly studied components of cortical and subcortical LFP’s related to movement (Buzsáki & Draguhn, 2004): theta (4-7Hz), alpha (8-12Hz), Beta (13-30Hz), and Gamma (60-90Hz). The second method identified “personalized” power bands, determined using a signal decomposition approach aimed at capturing the primary neural signals at each recording location.

The personalization approach first transformed time-domain LFP into a spectral representation using short-time fourier transform (STFT), then performed principal components analysis (PCA) on the z-scored STFT outputs to identify bands of neural activity capturing substantial spectral variance. A peak-finding algorithm was finally applied to the PCA component weights to find the frequency range best approximating each PC (Figure 3A). This process was run on an independent, multi-hour dataset. The resulting bands often overlapped canonically described power bands (Figure 3B). The top two PC’s in all cortical regions reflected personalized versions of beta and gamma, capturing 11-17% and 7-9% of the overall spectral variance, respectively. The top two PC’s in the STN were found in low-beta and gamma ranges, with gamma (11-31%) accounting for a larger proportion of the spectral variance than beta (5-7%). The third and fourth PC’s were more variable across brain areas, typically representing a combined theta-alpha band or low-gamma.

**Figure 3.**
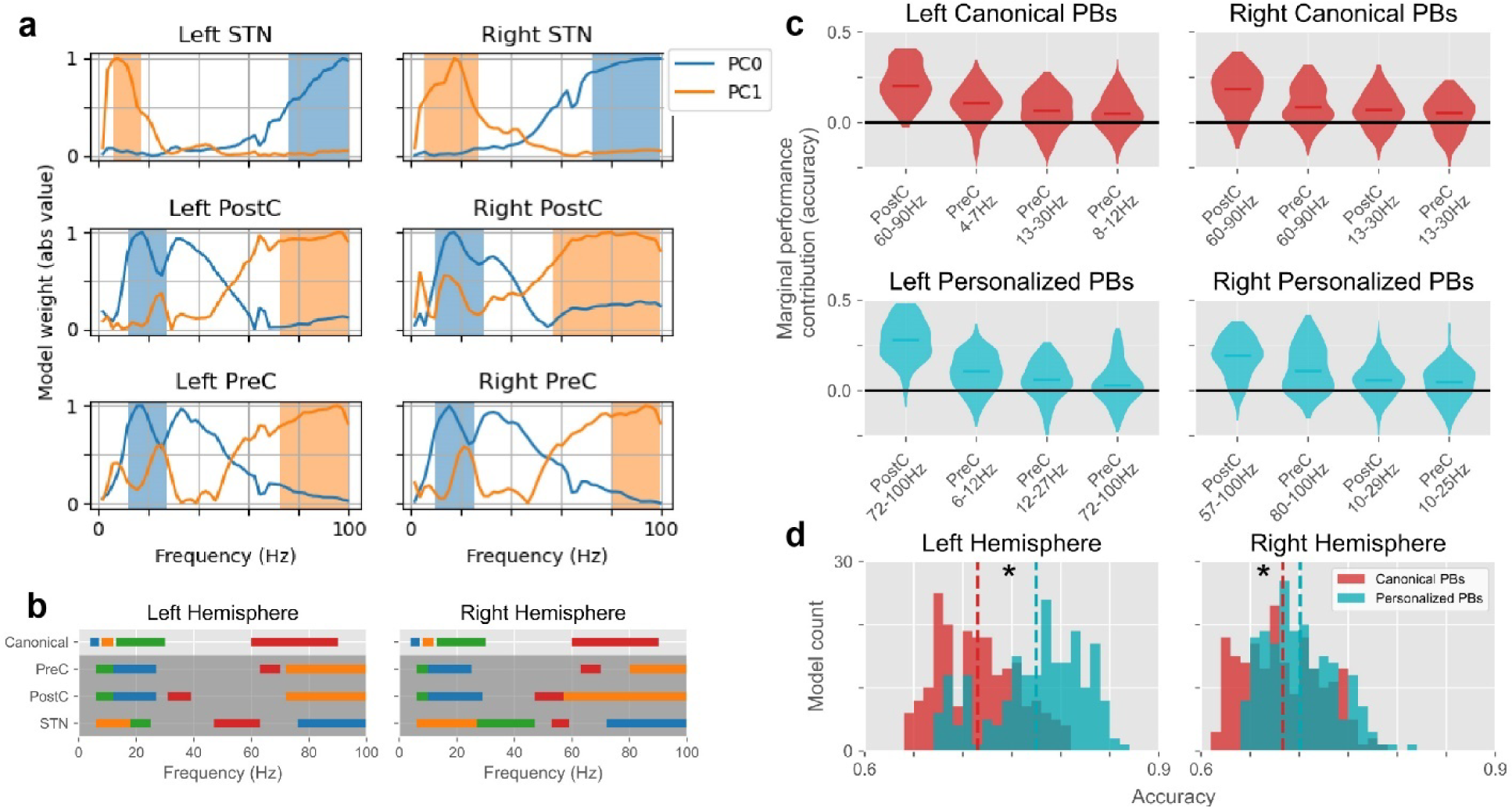
Personalizing power bands improves algorithm performance. (a) To obtain Personalized power bands, principal component (PC) models were fit to spectrogram data from an independent dataset where the participant was freely behaving in their home for 2 hours. The PC weights (displayed for the top two PC’s of each recording channel) were used with a peak-finding algorithm to identify contiguous frequency ranges that approximated each PC in a way that was compatible with the device-embedded controller. Note that frequencies below 4Hz and above 100Hz were excluded due to known noise issues. (b) The top four personalized power bands were selected for each of the three recording locations, resulting in 12 unique biomarkers per hemisphere. The frequency ranges for each Personalized power band are displayed for comparison with Canonical power bands, with the color indicating the PC number (blue: PC0, orange: PC1, green: PC2, red: PC3; colors arbitrary for the canonical bands). (c) The marginal performance contribution for the top four power bands (ranked by mean, indicated by horizontal lines) are displayed for each power band pool. (d) The top 200 models from each of the biomarker pools (Canonical and Personalized) were directly compared to determine if the Personalized power bands provided significant performance benefits as compared to the Canonical power bands. This performance difference relates only to the choice of input power bands. Parameter optimization was performed identically for all models from both pools, therefore isolating the additive impact of power band personalization on top of the performance gains from the rest of the machine learning pipeline. Vertical dotted lines indicate the mean of each distribution. The top Personalized power band models outperformed their matched-rank Canonical models by an average of 6% (left hemisphere) and 2% (right). Asterisks indicate statistical significance at p<0.001 (permutation test - difference in means).

All combinations of power bands from the candidate pools were considered during feature selection. For each of the two candidate pools, combining all three brain regions, four power bands in each region, and at most four total inputs yields 682 models. These were narrowed down to three candidates per pool by selecting the models with the highest F1 scores after 20 iterations of Bayesian optimization (“Broad search”, Figure 2C). The models that advanced from the broad search were further optimized with an additional 200 iterations (“Final optimization”, Figure 2C), and the single top performing model was then selected for final analysis and aDBS experiments.

During optimization, we investigated whether inclusion of cortical recordings and power band personalization added value to the algorithm. We first computed the marginal performance contribution of each individual power band as a measure of feature importance using classifier accuracies from the broad search. The top power bands were uniformly located in cortical locations, indicating that ECoG signals were most valuable for decoding movement (Figure 3C). We then directly compared the two power band identification methods to assess the personalization procedure. Accuracy for the top personalized model was 6% greater than the top canonical model for the left hemisphere, and 3% greater for the right. For both hemispheres, this effect was consistent when comparing the top 200 models from each pool, displaying significantly greater mean accuracy for the personalized bands (p_left_,p_right_<0.001, permutation test; Figure 3D). Personalized power bands therefore offered significant performance improvements and were selected for subsequent analysis and adaptive therapy.

### Evaluation of offline movement classification

We evaluated performance of the final neural classifiers using a single held-out session to determine whether they would be feasible for guiding adaptive stimulation (Figure 4A). Prediction accuracy for the left and right hemispheres was 83% and 76%, respectively (binomial test H_0_: accuracy=0.5, p_left_,p_right_<0.001). Both True Positive Rate (TPR, 88% and 93%) and True Negative Rate (TNR, 81% and 68%) were significantly above chance (binomial test H_0_: TPR, TNR=0.5, p_left_TPR_,p_right_TPR,_p_left_TNR_,p_right_TNR_<0.001; Figure 4B), yet there remained modest prediction bias towards the moving state (8% and 18%). Subthreshold acceleration of the inactive arm occasionally occurred during unimanual arm movements and was predicted by the classifier as contralateral movement (Figure 4A, red arrow), contributing to this bias. Accelerometry distributions segregated according to predicted movement state (Figure 4C), illustrating the classifier’s ability to discern true movements. Finally, moment-by-moment stimulation amplitudes were simulated (ramp rates: 1.6mA/sec; state targets: 1.6-2.2mA) to predict neurostimulator activity in later adaptive experiments (Figure 4D). As intended, stimulation was consistently high in each hemisphere during contralateral arm movements (left: 86% of the time, right: 90%), but not when stationary (left: 17%, right: 31%).

**Figure 4.**
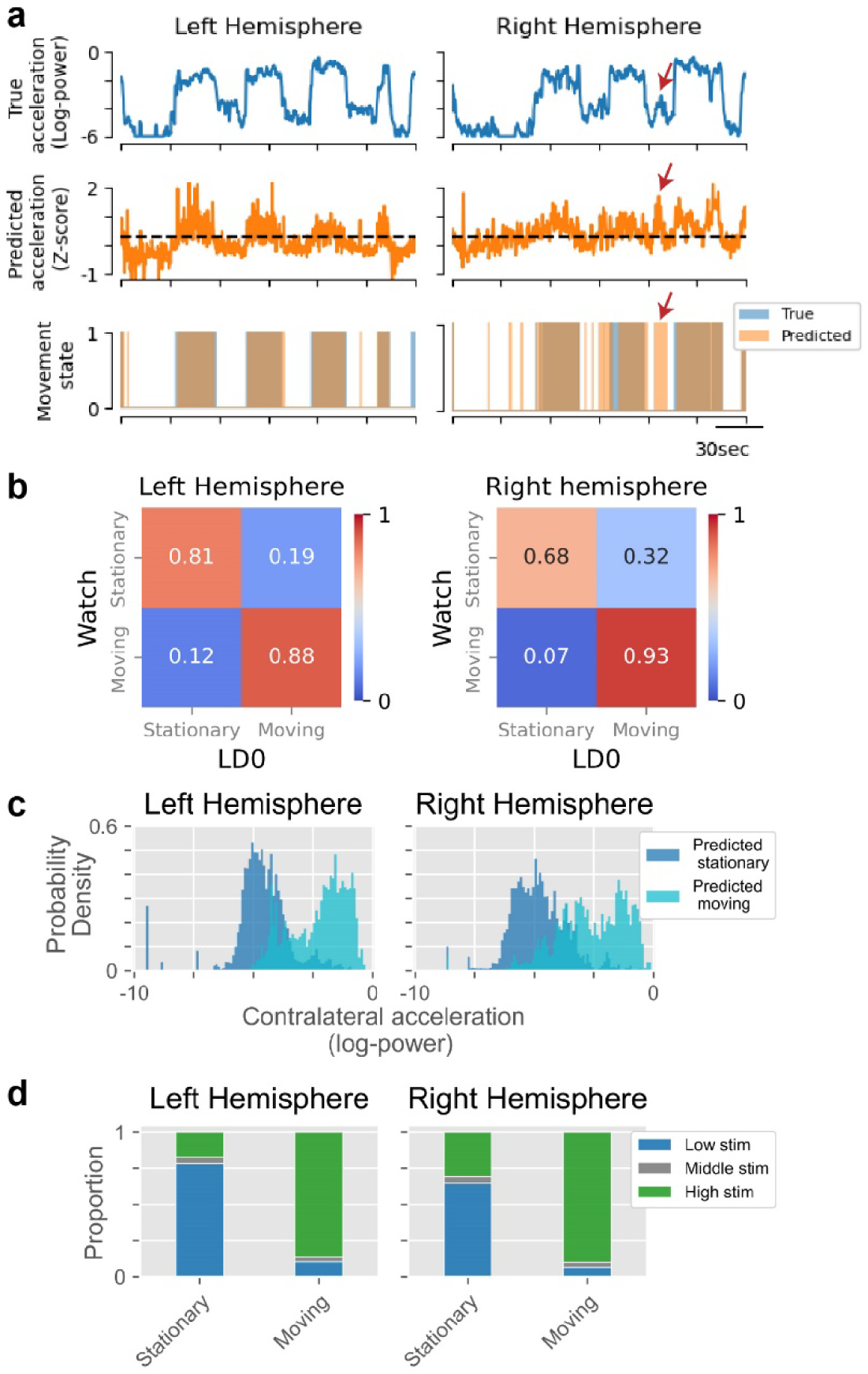
Final offline model performance. (a) An example time window of decoder predictions is displayed with the true acceleration (top row), the predicted acceleration (middle row) and an overlay of the true (blue) and predicted (orange) movement states (bottom row). A notable false positive is indicated with red arrows on the right hemisphere plots, highlighting a common observation: even when the neural classifier incorrectly predicted movement, there was often observable sub-threshold acceleration. (b) Confusion matrices are displayed for decoder predictions from a held-out dataset. (c) Distributions of the accelerometry during predicted movement are compared to predicted stationary periods to further assess the discrimination performance of the neural decoder. Acceleration of the left hand (right hemisphere) notably lacked strong bimodality, making binary movement state predictions challenging. (d) The moment-by-moment stimulation current was simulated according to the predictions of the neural classifier and the stimulation parameters that were used in the later online adaptive experiments. The proportion of time spent at each stimulation amplitude is plotted for each true movement state of the offline test data.

### Evaluation of movement classification during aDBS

We compared two aDBS algorithms with constant DBS for evaluating the therapeutic impact of movement-responsive stimulation (Figure 5A). The “Movement Responsive” condition consisted of increasing stimulation during predicted movement, while the “Inverted” condition had the inverse association, decreasing stimulation during predicted movement. The Inverted condition, predicted to have the opposite impact of the Movement Responsive condition, acted as a control for the possibility that stimulation intermittency was the driving factor of benefit. Both aDBS conditions varied stimulation between low (1.6mA) and high (2.2mA) levels, titrated prior to experimentation as tolerable for the participant. The third “Constant” condition provided conventional therapy at an intermediate level (1.9mA) regardless of classifier predictions. Data were collected for each of the three conditions in every session using a randomized and patient-blinded ordering. The same movement tasks that were completed during algorithm training were again performed in each block of the aDBS experiments, except for hand open-close, which was replaced with nose-tapping to represent ballistic arm movements involving proximal musculature. 12 sessions of data were collected over a 154 day period, with the last day performed 435 days after the original training data were collected.

**Figure 5.**
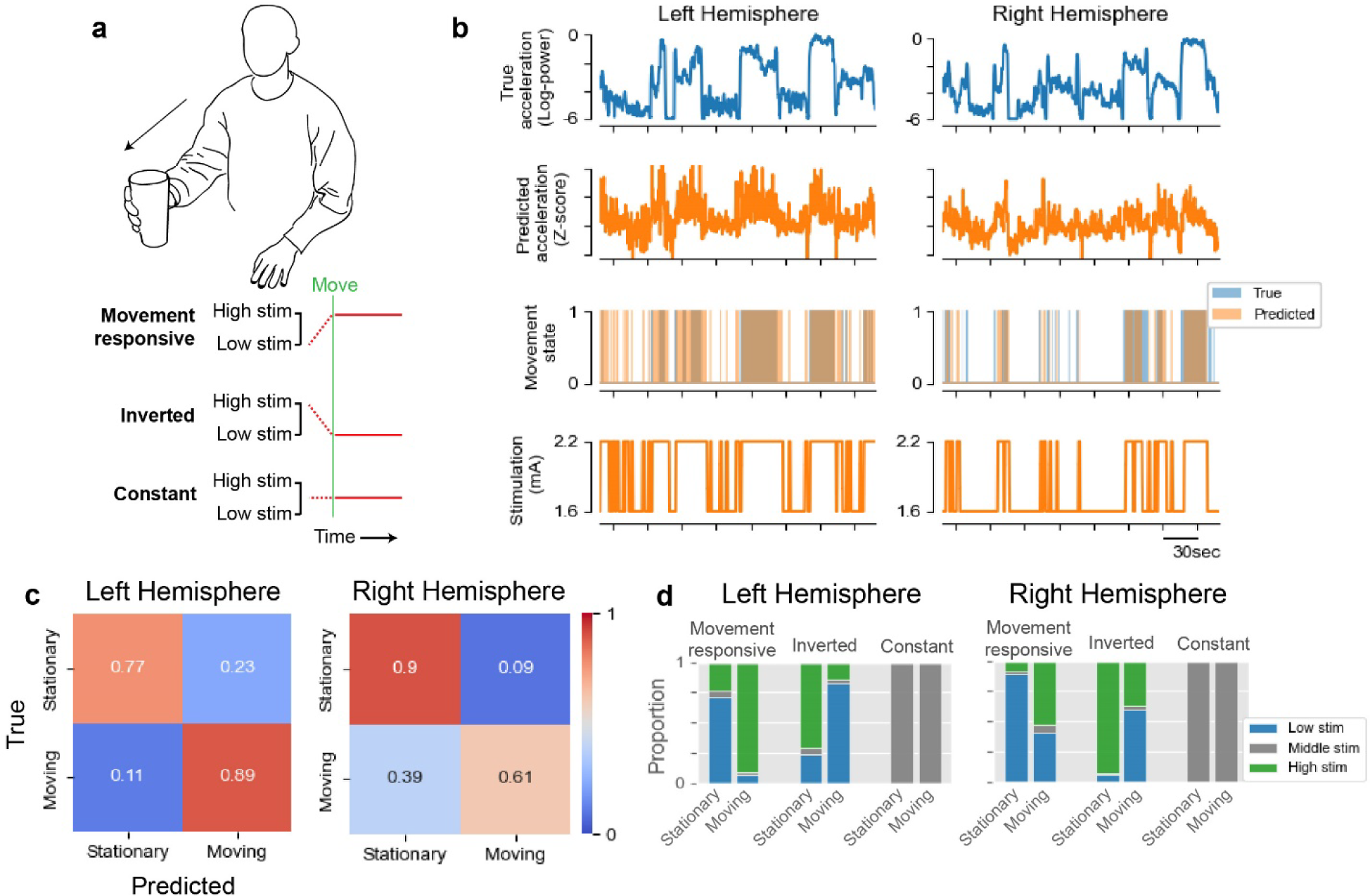
Performance of neural classifiers is maintained during online adaptive stimulation. (a) For evaluating movement-responsive DBS, three test conditions were used: “Movement responsive” increased stimulation during sensed movement, “Inverted” decreased stimulation during sensed movement, and “Constant” kept stimulation at an intermediate level. (b) Example data from the Movement responsive block of a single session displays the wearable accelerometry (top row) and neural predictions of the accelerometry (second row) together with the corresponding discrete movement states (third row) and the responsive stimulation adjustments (bottom row). (c) Confusion matrices depict the state-specific prediction accuracy from the neural classifier averaged across conditions. (d) The proportion of time at the low, high, and intermediate stimulation amplitudes was computed separately for each movement state and test condition. The goal of the algorithm was to maximize the time spent at high stim when in the moving state, while minimizing the time spent at high stim when in the stationary state (and vice-versa for the Inverted condition).

The movement classifier continued to perform well during aDBS (Figure 5B, Supplementary Video 1). Performance was stable for many months at a time; however, changes in neural signal magnitude were observed on two occasions more than a year after training data collection (once in each hemisphere, days 384 and 429). These were addressed through manual adjustment of algorithm thresholds without modifying other parameters, which restored performance to previous levels (Supplementary Figure 1). Prediction accuracy for the left and right hemispheres across all testing sessions was 82% and 76%, respectively (binomial test H_0_: accuracy=0.5, p_left_,p_right_<0.001), and did not differ substantially across stimulation conditions (Supplementary Figure 2). The classifiers displayed mild prediction bias. An 8% bias towards predicting movement was observed for the left hemisphere, while a 14% bias towards no movement was observed for the right (Figure 5C). Overall, intended stimulation contingencies were achieved for each condition (Figure 5D).

### Self-perceived therapeutic effects

We first obtained subjective therapeutic assessments by asking the participant to blindly score each of the blocks on perceived quality of movement (Figure 6A). One-way ANCOVA with performance order as a covariate revealed a significant effect of condition on self-scores (p=3.1e-4). Pairwise T-tests showed significantly reduced self-scores for the Inverted condition as compared to both the Movement Responsive (p=5.7e-4) and Constant (p=1.9e-4) conditions. Adjusting stimulation with respect to movement state therefore influenced subjective therapeutic quality, though primarily driven by reduction in quality reported during the Inverted condition.

**Figure 6.**
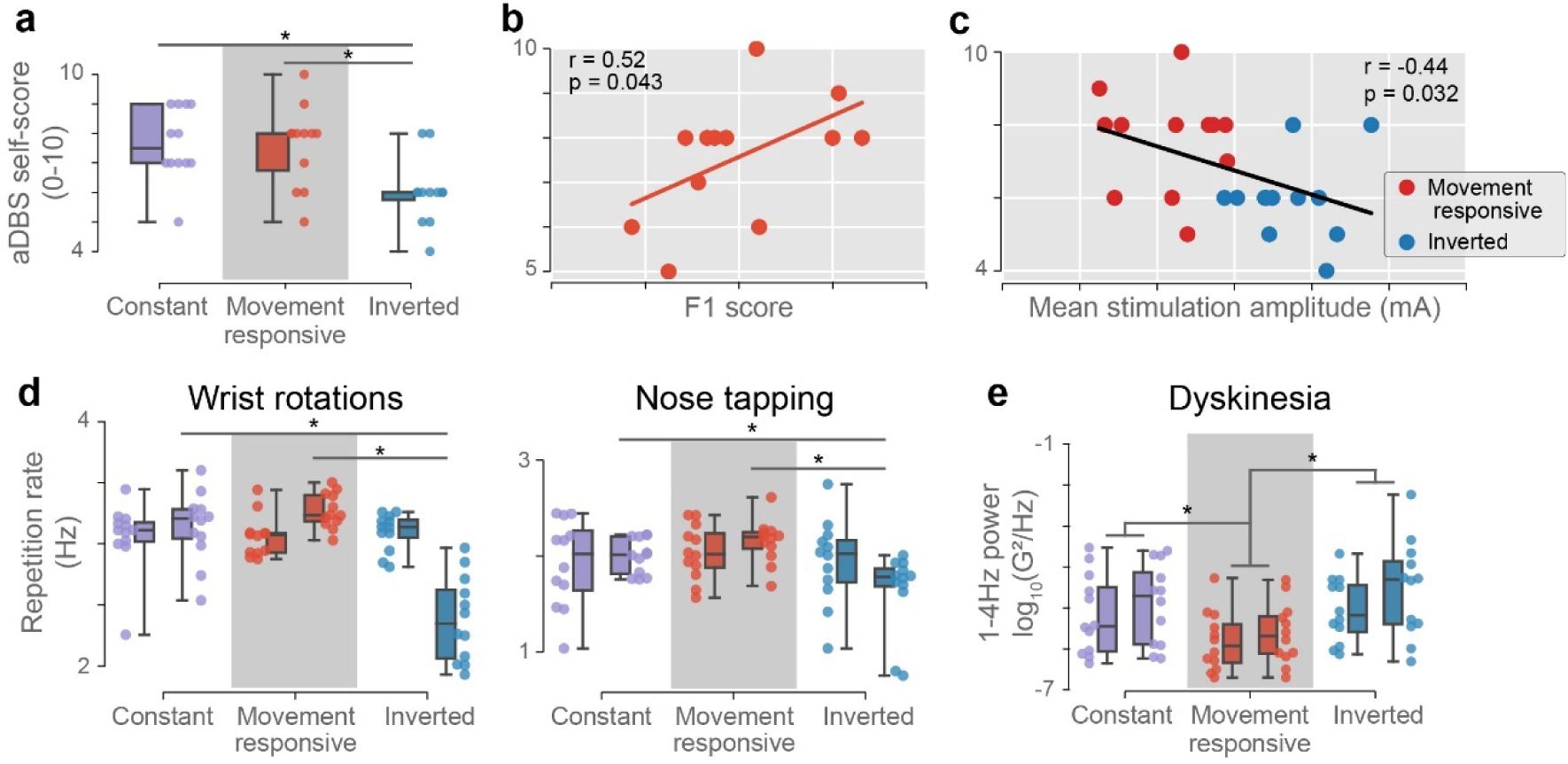
Movement responsive stimulation impacts self-perceived therapeutic quality, movement speed, and dyskinesia in an accuracy-dependent manner. (a) Blinded self-scores were reported by the subject at the conclusion of all 12 sessions to quantify the participant’s self-perceived therapeutic quality of each stimulation condition. The order in which the conditions were performed in each session was randomized in balanced fashion and was included as a covariate in all across-condition statistical testing. (b) The impact of classifier performance on therapeutic outcomes was assessed by regressing self-scores on the F1 score on a per-session basis. A single self-score was provided for both hands; the reported F1 score represents the average over the two hemispheric classifiers. Only data from the Movement responsive condition are shown. (c) As a control analysis to determine if therapeutic outcomes were simply better with higher average stimulation (e.g., from classifier biases), self-scores were regressed on mean stimulation amplitude across the two hemispheric stimulators on a per-session basis. (d-e) Quantifiable features of movement from representative motor tasks are shown independently for each hand, with the left bar and datapoints in each pair representing the left hand. (d) Movement speed was assessed using the repetition rate during two repetitive motor tasks: wrist rotations (left) and nose-tapping (right). Note that the right hemisphere (left hand) classifier displayed a bias towards predicting stationarity during movement. (e) Dyskinesia was assessed during the bimanual rest period where movement was assumed unintentional. Low-frequency power in the wrist-worn accelerometry data was used to quantify dyskinesia. Note that both hemispheric classifiers were highly accurate at predicting the stationary state. P-values were determined by one-way (a) and two-way (d-e) ANCOVA, and Pearson correlation (b-c). Asterisks indicate statistical significance at p<0.05. All box plots display mean, interquartile range, minimum, and maximum.

Furthermore, we sought to determine if self-scores depended upon algorithm performance or other confounding factors. A positive correlation between self-scores and classification performance is predicted under the foundational hypothesis that increasing stimulation during movement provides therapeutic benefit. We indeed observed a significant correlation between classifier performance and self-scores across sessions (Figure 6B; Pearson r=0.52, one-sided p=0.043). Additionally, we investigated whether greater average stimulation was simply driving perceived differences in therapeutic quality for the two aDBS conditions. Counter to this explanation, a significant negative correlation was observed between mean stimulation amplitude and self-score (Figure 6C; Pearson r=-0.44, two-sided p=0.032). Collectively, these results suggest that self-perceived motor improvements resulted from accurately targeting movement states and not indiscriminately increasing stimulation (e.g. from classification biases).

### Movement speed and stimulation side-effects

We next tested the prediction that movement-locked increases in stimulation would enable faster movements, and stimulation decreases would slow movement. We quantified movement speed as the repetition rate for wrist pronation-supination and nose-tapping (Figure 6D). Two-way ANCOVA’s were performed with performance order as a covariate, revealing significant main effects of condition (p=1.3e-8), and hand (p=9.2e-3) for wrist rotations, and condition for nose-tapping (p=1.3e-3). A significant interaction between hand and condition was observed for both wrist rotations (p=1.1e-9) and nose-tapping (p=0.019). Pairwise T-tests showed that, for both movement types, speeds were slower in the Inverted condition compared to other stimulation conditions for the right hand only (p<0.01). This suggests that movement speed was modulated by stimulation condition for the right dominant hand alone. Importantly, classification of the moving state specifically for the left non-dominant hand was lower than any other state predictions (Figure 5C, Right Hemisphere). This resulted in stimulation being lower on average during left hand movements as compared to the right. However, stimulation was still consistently low during rest for both.

We further predicted that dyskinesia, a common side-effect of dopaminergic medication and DBS (Maciel et al., 2021), would be reduced during rest periods of the Movement Responsive condition when stimulation amplitudes were low. We quantified dyskinesia using low-frequency power (1-4 Hz) of the watch accelerometry (Rodríguez-Molinero et al., 2019; Figure 6E). Two-way ANCOVA with performance order as a covariate revealed significant main effects of condition (p=1.0e-6), and hand (p=0.018), but no significant interaction (p=0.61). Data for the two hands were therefore grouped for pairwise T-tests that showed significantly reduced dyskinesia in the Movement Responsive condition compared to other stimulation conditions (p_vs_constant_=8.2e-6, p_vs_inverted_=1.1e-6). We also evaluated tremor, quantified using accelerometry power in the 4-7 Hz range (Dai et al., 2015), which was previously identified as containing tremor markers for this participant at low stimulation (data not shown). We found no evidence that Movement Responsive aDBS worsened tremor, as power in the 4-7 Hz band was also reduced (Supplementary Figure 3). In summary, Movement Responsive stimulation provided a bilateral reduction in resting state dyskinesia, and a unilateral increase in movement speed impacting the dominant hand.

### Naturalistic typing performance

We finally investigated movement quality in a bimanual setting by analyzing typing performance, predicting that movement-locked stimulation increases would enable faster action without compromising dexterity. Three metrics were analyzed (Figure 7): mean keypress duration (quickness of individual finger movements), overall typing speed, and backspace rate (error rate). For keypress duration, one-way ANCOVA with performance order as a covariate revealed a significant effect of condition (p=4.6e-7). Pairwise T-tests showed significantly shorter keypress durations for the Movement Responsive condition as compared to both other conditions (p_vs_cconstant_=8.2e-6, p_vs_inverted_=1.1e-6). For typing speed, a one-way ANCOVA again revealed a significant effect of condition (p=0.031). A significant pairwise difference was observed between the Movement Responsive and Inverted conditions (p=8.5e-3), with Movement Responsive typing being faster by 0.33 keypresses/s on average. No significant differences were observed for backspace rate (ANCOVA, p=0.092). Movement-responsive stimulation therefore impacted both quickness of individual finger movements and overall typing speed as predicted without significantly compromising dexterity.

**Figure 7.**
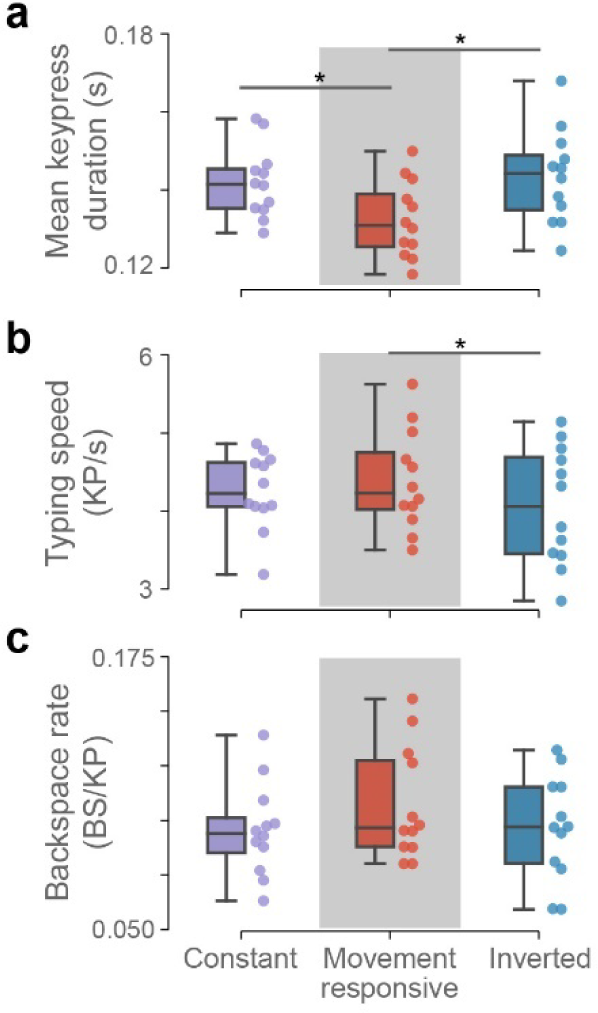
Movement adaptive stimulation increases keypress responsiveness during naturalistic typing. Three metrics of typing performance were evaluated across stimulation conditions. (a) Movement responsive stimulation resulted in a significant decrease in mean keypress duration relative to both of the other conditions. (b) Faster individual keypresses translated to an overall increase in typing speed when comparing only the Movement Responsive and Inverted conditions. (c) No significant differences in backspace rate (errors) were observed. KP: keypress; BS: backspace. Asterisks indicate statistical significance at p<0.05 (one-way ANCOVA). All box plots display mean, interquartile range, minimum, and maximum.

## DISCUSSION

The dynamic nature of PD suggests that adaptive stimulation may be a better treatment option than constant stimulation. The varied timescales and sources of these dynamics have given rise to many proposed aDBS paradigms that fit broadly into three categories: algorithms that respond rapidly to specific phasic or transient brain signals for manipulating low-level circuit behavior (Grado et al., 2018; He et al., 2023; Holt et al., 2016; Little et al., 2013, 2016; Popovych & Tass, 2019; Tinkhauser et al., 2017), algorithms that use neural biomarkers to track gradual changes in PD clinical states (Arlotti et al., 2018; Gilron, Little, Perrone, et al., 2021; Oehrn et al., 2023; Rosa et al., 2017; Swann et al., 2018), and algorithms that tune stimulation to the current behavioral setting (e.g., movement (Darbin et al., 2022) and sleep (Gilron, Little, Wilt, et al., 2021; Smyth et al., 2023)). This study fits into the third category, matching the prokinetic strength of DBS to moment-by-moment motor demands. However, this feedforward approach can only work if the effects of DBS materialize quickly enough to impact movement. Critically, we have shown that quantifiable alterations in motor performance were discernible on a rapid timescale (Figure 6), with movement epochs lasting only 20 seconds. This is notable given that STN-DBS effects during simple on-off testing typically operate on the scale of seconds to hours (Hristova et al., 2000; Koeglsperger et al., 2019). These results suggest that movement-responsive stimulation can strengthen therapeutic efficacy while minimizing side-effects through rapid behavior-dependent switching.

Importantly, effective use of movement-responsive aDBS was found to depend on accuracy of the classifier (Figure 6B). Two key techniques were used to improve this accuracy. First, we employed a power band personalization method that was superior to using canonical power bands (Figure 3D). Second, we included ECoG signals, which were found to be the most valuable for decoding during our feature selection routine (Figure 3C). This adds to a growing body of support for the inclusion of ECoG sensors in aDBS systems (Gilron, Little, Perrone, et al., 2021; Merk et al., 2021; Oehrn et al., 2023). Stability of these signals is also critical to maintain performance. ECoG is considered a stable signal for brain-computer interfaces (BCI) (Degenhart et al., 2016; Volkova et al., 2019), and has been shown robust for decoding over multiple years (Blakely et al., 2009; Pels et al., 2019). In our study, scaling of the ECoG signals was observed on two occasions (once per hemisphere) more than a year after initial model training, and was addressed by manually adjusting algorithm thresholds (Supplementary Figure 1). While the source of signal disturbance is uncertain, stability before and after manual adjustments suggests that typical inflammatory responses were not the cause (Degenhart et al., 2016). Minor electrode position changes could be the cause, as this study involved a more mobile, unconstrained deployment compared to previous investigations of ECoG-based BCI’s. Nonetheless, the isolated effect of signal scaling may be readily accounted for with auto-scaling, and the overall performance of both decoders was encouraging for long-term use. Future work should analyze this stability more thoroughly and explore adaptive re-scaling protocols.

Several other avenues for continued research and engineering improvements are motivated by the foundational results of this report. We established that movement-responsive stimulation has a positive impact on movement (Figure 6-7), yet the extent of that clinical impact was here limited by the experimental design requirement to counterbalance the two aDBS conditions (to match average stimulation amplitudes). Any improvement from the primary condition was limited by how much reduction in efficacy could be tolerated in the Inverted control condition while preserving patient comfort. Future studies should create an unconstrained Movement Responsive stimulation contingency, beginning with more pronounced stimulation level differences. Graded stimulation proportional to the intended movement vigor should also be explored (Meidahl et al., 2017), since movement is not truly binary in most contexts. Additionally, coordinating action of the two stimulators may offer opportunity for improvement, since unilateral DBS is known to impact symptoms bilaterally (Shemisa et al., 2011). Finally, this approach, validated in PD, could provide benefit in other conditions that are impacted by reductions in movement. Specifically, this technique could benefit patients with cerebrovascular injuries (stroke), where there is a reduction in motor initiation and vigor. The present study has focused on the acute benefits of movement-responsive DBS augmenting movement as an assistive BCI. However, this approach could also support the selective facilitation of network learning and plasticity if used as a rehabilitative BCI tool though motor imagery combined with neurofeedback. This could be used to preferentially reinforce high gain movements (Yttri & Dudman, 2016) by directly linking cortical motor initiation signals to basal ganglia stimulation, and therefore potentially be more effective than current peripheral rehabilitative BCI, dependent on virtual reality, robotics or direct muscle stimulation.

On a closing technical note, this study introduces a framework for aDBS optimization with three key pillars that should inform future aDBS research and technology development. First, to build algorithms that work well at home, data must be collected naturalistically, as neural coding changes across laboratory and naturalistic settings (Jackson et al., 2007; Wang et al., 2016; Wu et al., 2006). Notably, our training pipeline is compatible with simpler hardware implementations, including only wrist-worn accelerometers, to facilitate naturalistic deployment. Second, to perform data-driven optimization of a device-embedded algorithm, one must be able to simulate its operations. This was achieved using the *rcssim* software package, a high-fidelity *in-silico* device simulator developed in support of this study. Third, a tailored machine learning pipeline is required for optimizing aDBS performance. While the elementary operations of the RC+S device are simple, their interactions become highly complex, making conventional optimization approaches impossible. The future of aDBS therapies will rely upon such frameworks to optimize programming in a tractable way.

## CONCLUSION

We demonstrate for the first time the clinical efficacy of a fully embedded movement-responsive aDBS algorithm. The algorithm, which was remotely optimized in the participant’s own home, improved self-reported scores of therapeutic efficacy as well as movement speed, dyskinesia, and typing performance. These findings provide early support for aDBS algorithms that modulate stimulation in accordance with movement state for the treatment of PD and address challenges related to automated parameterization supporting scalable aDBS.

## METHODS

### DBS implant and patient information

This study was reviewed by the University of California, San Francisco Institutional Review Board and registered on clinicaltrials.gov (IDE G180097; NCT03582891). One participant with PD was recruited from a parent trial testing slow aDBS and enrolled in the study after providing written informed consent. The participant had been previously implanted with one pair of electrode leads in each hemisphere, consisting of (A) quadripolar depth electrodes (Medtronic model 3389) in the STN, and (B) a quadripolar subdural ECoG array (Medtronic model 0913025) spanning the arm/hand regions of the precentral and postcentral gyri (motor and somatosensory areas). Each pair of electrode leads was connected to an investigational sensing-enabled INS (Summit RC+S, Medtronic) in the ipsilateral subclavicular space. The INS provided therapeutic stimulation to the STN through the depth electrodes, recorded LFP signals from the depth and ECoG electrodes, and recorded accelerometry data through an accelerometer in the INS case. All stimulation changes were self-initiated by the participant, and participant safety was ensured by allowing them to revert to their standard, pre-programmed DBS settings at any time using a handheld patient programmer device.

### Neural data sensing configuration and signal processing

All neural recordings were performed during monopolar stimulation. Bipolar recordings in the STN were collected using a “sandwich” configuration (electrodes on either side of the stimulating contact) for common-mode rejection of the stimulation artifact. Two channels of neural data were recorded along the ECoG array, using the first and second electrodes for one channel (postcentral gyrus) and the third and fourth electrodes for the other (precentral gyrus). Neural time-domain data were recorded at a sampling rate of 500Hz. Device-embedded signal processing consisted of short-time Fourier transforms (STFT) with an FFT size of 256 pt and an interval of 50 ms.

### Behavioral data streaming, signal processing, and synchronization

Detailed descriptions of the behavioral data collection platform can be found in Strandquist et al., 2023. Briefly, arm movement data were collected using Apple Watches (Apple, Inc) worn on each wrist and uploaded to a remotely accessible third-party data repository (Rune Labs, Inc.). These data were sampled at 50 Hz, and power was calculated using STFT with an FFT size of 64 pt and interval of 100 ms. Power was calculated in the 0-5 Hz range for movement decoding, 1-4 Hz for quantifying dyskinesia (Rodríguez-Molinero et al., 2019), and 4-7 Hz for quantifying tremor (Dai et al., 2015). Video data for participant observation and kinematic pose estimation were recorded on three cameras arranged at varied angles in the participant’s home office. A custom keylogging application was built to provide statistics regarding typing performance without compromising privacy by removing the letter identity of each keypress. This application was deployed on the participant’s home office computer with patient knowledge and consent. All data was securely transferred to remotely accessible databases over a virtual private network (VPN). To overcome drift and time discrepancies across system clocks and align the data streams, a synchronization event was performed at the start of all recordings. The participant tapped each side of their chest over the implanted INS while standing in view of the cameras. This movement created identifiable spikes in the INS and Apple Watch accelerometry signals and could be discerned on the video, allowing realignment of all data to a common time.

### Data collection protocols

Three different datasets were collected remotely for this study. For identifying personalized power band candidates, a single two hour session of free behavior was collected during monopolar stimulation at 1.6 mA. The participant was instructed to go about their regular daily routine while occasionally taking breaks to perform hand open-close movements and rest their hands on their lap. Since this dataset was only used to identify the most prominent components of the neural signals through PCA, there were not firm constraints on the participant’s actions during data collection other than ensuring that there were periods of both movement and non-movement. Separate datasets were used for identifying the personalized power bands and evaluating their use in decoding movement to ensure that we avoided any risk of overfitting and represented generalizable performance.

A second dataset was collected for optimizing the movement classifier and performing feature selection from the identified power band candidates. This dataset included six days of data where the participant performed a prescribed set of motor tasks at three different levels of monopolar stimulation (1.2 mA, 2.4 mA, 2.6 mA). Each day consisted of approximately 25 minutes of total recording time. The participant was given an instruction sheet with the list of motor tasks, which included standardized movement tasks from the Unified Parkinson’s Disease Rating Scale Part III (UPDRS; sections 3.17-3.18 rest tremor, 3.4 finger tapping, 3.5 hand movements - open-close, 3.6 hand movements - pronation-supination) as well as blocks of text for typing that were each approximately 100 words long (Figure 2A,B). The participant was a proficient typer prior to beginning the study.

For evaluating the movement responsive aDBS algorithm, 12 days of data were collected while the participant performed a similar set of tasks in three different stimulation conditions (Movement-responsive, Inverted, Constant). Each day again consisted of approximately 25 minutes of total recording time. The open-close hand movements (UPDRS section 3.5) from the initial six day dataset were replaced with finger-to-nose tapping, and no movements of the lower body were performed. All movements were instructed to last 20 seconds, and the rest phase 10 seconds.

While optimal monopolar stimulation amplitudes were titrated to allow noise-free STN recordings with the sandwich sensing configuration, the participant’s standard therapeutic settings were conventional stimulation (cDBS) in a bipolar arrangement. In the monopolar setting, stimulation amplitudes were titrated separately for each of the three collected datasets before the first session. The high (2.2 mA) and low (1.6 mA) stimulation amplitudes for adaptive recordings were selected to be balanced around the constant stimulation amplitude (1.9 mA). These targets were also conservatively defined to ensure that the Inverted condition remained tolerable for the participant, since the inverted stimulation contingency was perceived as worse therapeutically. The main Movement Responsive test condition was therefore theoretically limited in how positive of an effect it could have, since its reciprocal needed to remain both counterbalanced and tolerable.

### Personalized power band identification and assessment

Personalized power bands were identified by performing Principal Component Analysis (PCA) on the power spectral data. A custom peak-finding algorithm was then used on each PC weight vector to approximate it with a single contiguous frequency range that was compatible with RC+S signal processing capabilities (Figure 3A). The resulting power bands therefore (A) aligned with the dimensions of the neural data that captured the most variance, and (B) produced features that were approximately linearly uncorrelated to facilitate downstream linear combination for classification models. The steps are briefly outlined below:

1. Compute a [n_samples x L/2] spectrogram of the time-domain neural data using the *rcssim* signal processing with the following parameters:

a. Fs_td = 500 Hz (time-domain sampling rate)
b. L = 256 (FFT size)
c. interval = 200 ms
2. Perform PCA on the z-scored spectrogram data and retain the top 4 PCs. Take the absolute values of each PC weight vector, which will be size [L/2 x 1] and represent the weight that a given PC places on each frequency bin in the previously performed FFT.
3. Run the peak-finding algorithm on the PC weight vectors to identify a contiguous range of frequency bins that best approximates each PC (restrict searchable range to 4-100Hz to avoid known noise issues):

a. Excluding frequency bins that have been included in power bands that were identified prior, find the frequency bin with the largest weight for the current PC (i.e., the peak of the weight vector)
b. Extend the current power band away from the peak in both directions until one of the following conditions are met:

i. The frequency bin has been included in a previously identified power band (prevents power band overlap)
ii. The weight on the frequency bin drops below 50% of the peak weight (excludes frequencies that are not well-represented in the PC)
iii. A trough has been encountered (prevents multi-peak power bands)
iv. The frequency bin is more than 25Hz from the peak (limits power bands to 50Hz full range)
c. Repeat for a total of 4 PC’s, independently for each anatomical recording area and brain hemisphere

Feature importance was assessed for each power band using its marginal performance contribution, which was computed as the performance increase (or decrease, as a result of overfitting) when a given power band was included as an additional feature in a model. Since models using every possible combination of four or less power bands were independently optimized in Step 1 of the feature selection process, the marginal performance contribution for a given power band was represented as a distribution of 196 values, one for each combination that included that power band.

### Movement classifier and optimization pipeline

The Summit RC+S embedded aDBS has four primary components to its onboard operations. (1) Neural signals are first processed into power band outputs given a set of sensing parameters that include the frequency edges dictating each band. (2) Up to four power bands are streamed, averaged across a number of subsequent samples dictated by the update rate, and linearly combined via feature weights into a continuous valued linear discriminant output (LD). (3) LD outputs are then passed through a series of logic operations, which include comparison to a threshold value and requisite hold times above (onset duration) or below (termination duration) that value to trigger a state change. (4) Upon state change, stimulation is ramped towards the target amplitude associated with the new state. Note that this is a non-exhaustive description of the device capabilities that is focused on the most important features and parameters for the present study.

Because of the unique operations of the RC+S, a custom optimization pipeline was required for training a movement classifier that leverages its full functionality. In brief, each loop of GPBO simulated the embedded algorithm for a given set of parameters and neural inputs, evaluated the algorithm performance against labeled movement data, and modeled the relationship between parameters and performance to support an intelligent and efficient search-based optimization. LD weights were the only parameters not modeled and optimized through the GPBO search method, but instead were fit by linearly regressing the continuous-valued wearable accelerometry on the neural signals within each GPBO loop. By fitting these weights in closed form rather than as additional parameters in the GPBO gaussian process model, the GPBO routine was greatly simplified. Notably, the thresholds were not fit in closed form (e.g., through linear discriminant analysis), since it was assumed that interactions between thresholds and parameters such as onset duration would require that they be jointly optimized. Using the regression technique, LD outputs therefore represented an estimate of the continuous-valued accelerometry, which was then processed through the remaining algorithmic steps to produce binary state outputs given the set of parameters defined in the current GPBO loop. These state outputs were then compared against the true movement state determined from wearable accelerometry to calculate an F1-score. The F1-score was used as the objective function of the GPBO search, which was selected as the performance metric to promote unbiased and accurate predictions. Prediction bias reported elsewhere in the results was calculated as the difference between the frequency of labels predicted by the classifier and the frequency of labels in the actual data. Optimization was performed using five-fold cross-validation, using a separate day of data for each fold. All optimization code was written in Python, using the *rcssim* package for simulating RC+S operations and *sklearn* and *skopt* for model fitting and optimization.

As a minor stage in this process, a state change blanking variable dictating a refractory period to allow stabilization of artifacts associated with changes in stimulation was also optimized. This simple step consisted of modeling the artifact, which was different at cortical and subthalamic recording locations, and automatically assigning the appropriate duration of blanking given the neural inputs present in the model (i.e., whether inputs were from the cortex or subthalamic nucleus).

In summary, the following sensing parameters were optimized for each hemisphere:

1. Four power bands (though only using three was found to be optimal in the right hemisphere), each consisting of:

a. Frequency lower limit
b. Frequency upper limit

And the following algorithmic parameters were optimized for each hemisphere:

1. LD weights (four total, one for each power band)
2. Update rate
3. Threshold
4. Onset duration
5. Termination duration
6. Lag (not a device-programmable parameter, but allowed for time delay between neural signals and movement)
7. State change blanking

In total, 16 sensing parameters and 20 algorithmic parameters were therefore optimized.

### Statistical testing

All statistical testing and data visualization was performed using the Python programming language and common statistical packages. Basic statistics were performed using *scipy*, and ANCOVA models were performed using *statsmodels*. Following ANCOVA, T-tests were performed as planned contrasts only in the event of statistically significant ANCOVA effects, thus no adjustments were made for multiple comparisons. The T-tests were covariate-adjusted for all statistically significant covariates identified through ANCOVA.

## Data availability

Anonymized data are available on the Dryad repository at https://doi.org/10.5061/dryad.4xgxd25hw.

## Code availability

All machine learning and data analysis code is available on GitHub at https://github.com/Weill-Neurohub-OPTiMaL/move-responsive-adbs. The *rcssim* package is available on GitHub at https://github.com/Weill-Neurohub-OPTiMaL/rcs-simulation.

## AUTHOR CONTRIBUTIONS

T.C.D., J.A.H, and S.J.L. conceived the study and designed the research. T.C.D. and S.R. scheduled and oversaw the experiments. T.C.D. and D.L. designed and programmed the optimization pipeline and related tools. G.S., A.Z., T.F., and R.B. designed the data collection infrastructure. T.C.D., G.S., A.Z., and R.B. analyzed the data. P.A.S. performed the implantation procedures. T.C.D. wrote the manuscript, and all other authors performed reviews. J.L.G, J.A.H, and S.L.J. acquired funding and supervised the study.

