## Supplemental figures for "Movement-responsive deep brain stimulation for Parkinson’s Disease using a remotely optimized neural decoder"

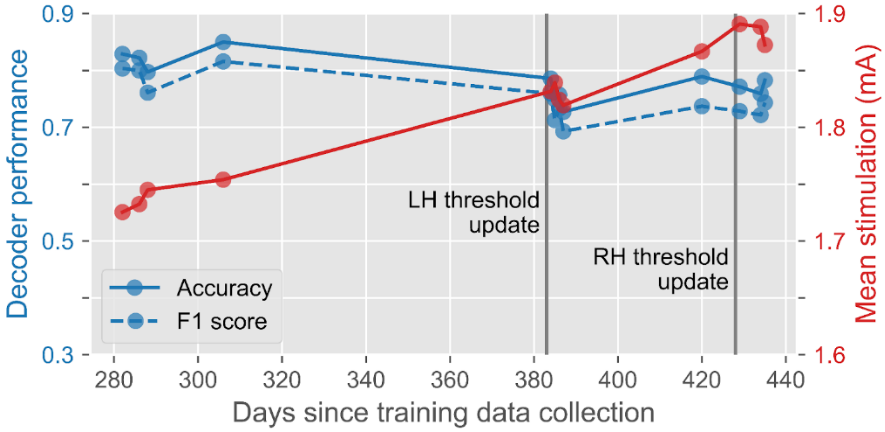


**Supplementary Figure 1. Decoder stability.**
Decoder performance and mean stimulation during the movement-responsive blocks are displayed. Each datapoint represents the average over the two hemispheres. Two manual updates were made to the threshold parameter of the algorithm due to sudden signal scaling: once for the left hemisphere on the fifth recording session 384 days after completion of the training data collection, and once for the right hemisphere on the tenth recording session 429 days after completion of the training data collection. Note that the mean stimulation is the result of both decoder biases and actual time spent in each movement state, and should therefore not be interpreted as a pure depiction of decoder performance or bias (refer to accuracy and F1 score for these two assessments, respectively).

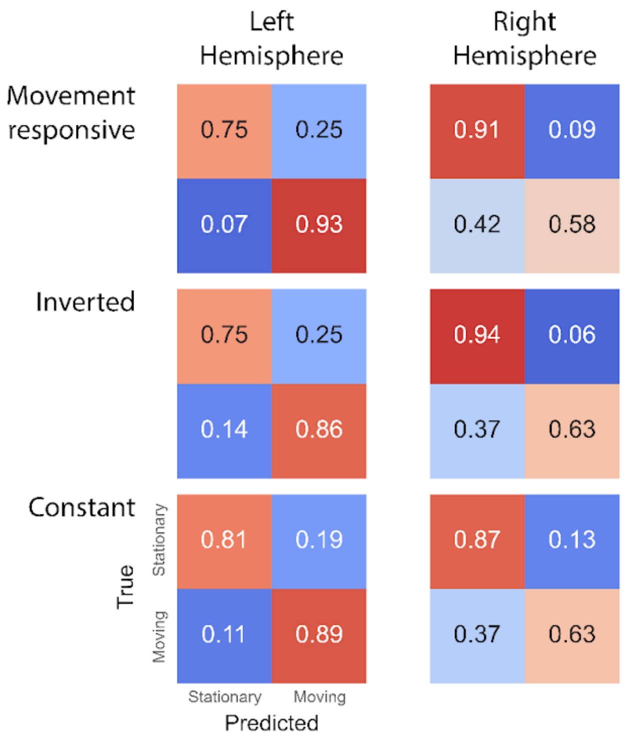


**Supplementary Figure 2. Condition-specific decoder performance.**
Confusion matrices showing decoder performance in each of the three stimulation conditions are displayed. Decoder performance did not vary substantially across the stimulation conditions despite their differential effects on behavior and stimulation artifacts.

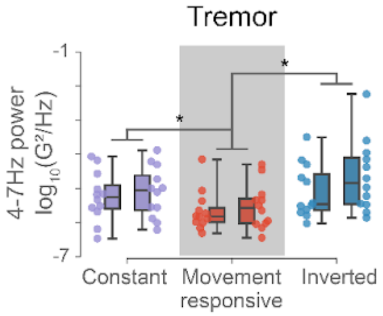


**Supplementary Figure 3. Effects of movement-responsive stimulation on resting tremor.**
Tremor was quantified using power in the 4-7Hz range of the wrist-worn accelerometers during rest behavior. The same results were observed as in the analysis of dyskinesia during the same periods (Fig. 6), with the Movement Responsive condition having less tremor than both other conditions. The observed trends likely reflect spectral bleed from the dyskinesia band (1-4Hz), as the movements appeared to be dyskinesia rather than tremor upon visual inspection of the video, and the on-average lower stimulation during rest in the Movement responsive condition would not be expected to ameliorate tremor.
